# Preoperative predictors of instrumental activities of daily living disability in older adults six months after knee arthroplasty

**DOI:** 10.1101/2022.07.09.22277442

**Authors:** Keigo Nanjo, Takashi Ikeda, Naoko Nagashio, Tomoko Sakai, Tetsuya Jinno

## Abstract

**Objective:** To investigate preoperative predictors of instrumental activities of daily living (IADL) disability in older adults six months after knee arthroplasty (KA).

**Design:** Prospective cohort study

**Setting:** General hospital with an orthopedic surgery department

**Participants:** Two hundred twenty (N = 220) patients 2: 65 years old with total knee arthroplasty (TKA) or unicompartmental knee arthroplasty (UKA)

Interventions: Not applicable

**Main Outcome Measure:** IADL status was evaluated for six activities. Participants chose “able,” “need help,” or “unable” according to their capacity of executing these IADL activities. If they chose “need help” or “unable” for one or more items, they were defined as “disabled.” Their usual gait speed (UGS), range of motion for the knee, isometric knee extension strength (IKES), pain status, depressive symptoms, pain catastrophizing, and self-efficacy were evaluated as predictors. Baseline and follow-up assessments were conducted one month before and six months after KA, respectively. Logistic regression analyses with IADL status as the dependent variable were performed at follow-up. All models were adjusted using age, sex, severity of the knee deformity, operation type (TKA or UKA), and preoperative IADL status as covariates.

**Results:** In total, 166 patients completed the follow-up assessment, and 83 (50.0%) reported IADL disability six months after KA. Preoperative UGS, IKES on the non-operated side, and self-efficacy were statistically significantly different between those with a disability at follow-up and those who did not and were therefore included in logistic regression models as independent variables. UGS (odds ratio = 3.22, 95% confidence interval = 1.38–7.56, *p* = 0.007) was determined as a significant independent variable.

**Conclusion:** The present study demonstrated the importance of evaluating preoperative gait speed to predict the presence of IADL disability in older adults six months after KA. Patients with poorer preoperative mobility should be provided careful postoperative care and treatment.

---

Knee osteoarthritis, a prevalent chronic disease related to aging, is increasing globally.^1,2^ Knee arthroplasty (KA), including total knee arthroplasty (TKA) and unicompartmental knee arthroplasty (UKA), is a well-established operative treatment for degenerative knee joint disease in patients suffering from pain and daily life disability. However, 10–20% of patients reportedly face subjective functional limitations after KA.^3–6^ Considering the increasing demand of KA, it is necessary to strengthen countermeasures against postoperative disabilities.

For the aging population, independence in performing activities of daily living is crucial. Basic activities of daily living (BADL) are fundamental skills required to independently care for oneself, such as toileting and dressing,^7^ whereas, instrumental activities of daily living (IADL) are more complex activities related to the ability to live independently, such as shopping, transportation, and performing household chores.^8^ Thus, IADL disability usually precedes that of the BADL.^9^ Various cohort studies have shown that disability in IADL can affect all-cause mortality,^10^ the rate of cognitive functional decline,^11^ and quality of life,^12^ having a substantial impact on geriatric mental and physical health.

Various preoperative factors have been reported as outcome predictors after KA.^13–16^ Psychological factors such as pain catastrophizing,^13,15,16^ depression,^13,15,16^ and self-efficacy^13^ have been identified as predictors of worse outcomes after KA. Preoperative rehabilitation focuses on improving muscle strength, knee range of motion, and mobility to improve postoperative outcomes.^17^ However, most studies defined disability as an impairment in both BADL and IADL. Considering the hierarchical relationship between IADL and BADL,^9^ determining the predictors of IADL disability in the geriatric population after KA is potentially useful for both preoperative and postoperative management. Multiple meta-analyses regarding postoperative rehabilitation revealed that most intervention periods had been set within six months after KA,^18,19^ meaning that disability should be addressed within a limited period. Additionally, it is desirable for patients with an elevated risk for IADL disability to be screened early, preferably before surgery.

Due to limited studies in older persons undergoing KA, the preoperative predictors for disability in IADL are unclear.^20–22^ This study investigated the preoperative predictors of IADL disability in older adults six months after KA. We hypothesized that certain motor functions, such as gait speed and psychological factors, might help predict IADL disability.

## Methods

### Participants

Participants who underwent unilateral TKA or UKA between May 17, 2019, and June 30, 2021, at Shonan Kamakura General Hospital were included in this study upon meeting the following inclusion criteria: (1) being ≥ 65 years of age and (2) diagnosed with knee osteoarthritis based on the guidelines^23^: a radiologic Kellgren–Lawrence score (KL-score) of ≥ 2 (definite osteophytes and narrowed joint space) and knee pain for > 3 months. Patients were excluded by the following exclusion criteria: (1) being diagnosed with post-traumatic osteoarthritis; (2) rheumatoid arthritis; (3) cognitive decline (mini-mental state examination score < 23); (4) serious pathologies, such as cancer and neurological fallout that could affect the test performance, e.g., muscle paralysis; (5) history of lower extremity operation except for KA; (6) preoperative BADL disability (Barthel index < 100); and (7) intention of having a KA on the other knee before our study’s assessment. A total of 321 patients were selected at the beginning of the study. Eighty-six of them were excluded because of: a history of lower extremity operation outside the knee joint region (n = 32), rheumatoid arthritis (n = 19), revision TKA or UKA (n = 18), Barthel index <100 (n = 7), mini-mental state examination score < 23 (n = 6), and post-traumatic knee osteoarthritis (n = 4). Of the 235 patients invited to participate, 15 refused, leaving 220 participants.

### Study design

This was a prospective cohort study conducted and reported using the STROBE guidelines.^24^ The institutional review board at Tokushukai Group Ethics Committee (No. TGE01198-024) approved the study design, and all participants gave written informed consent. This study was conducted following Declaration of Helsinki. All participants were assessed by physical therapists a month before the operation as a baseline measurement and six months after the operation as a follow-up measurement. One of five physiotherapists (with at least 5 years of experience and trained well before assessments) was randomly selected to conduct all assessments. Patients were permitted full weight bearing on postoperative day one and managed per routine clinical procedure. Patients started gait exercises with a walker cane on postoperative day one and a single cane on a postoperative day two. Ascending and descending stairs were practiced on postoperative day five. Patients who could independently walk with a single cane were discharged by postoperative days 10–16 for TKA and 5–12 for UKA. Baseline characteristics such as age, sex, body mass index, bilateral KL-score, Charlson comorbidity index,^25^ and intra-operative information were retrieved as potential covariates from clinical records.

### Measurements

#### IADL status

As proposed by Lawton and Brody, the six activity items (shopping, cooking, housekeeping, using transportation, doing laundry, and handling finances) were assessed on the IADL scale.^8^ Because handling finances includes commuting to the bank as part of the act, it was also assessed. Using a telephone and taking medications were excluded because knee osteoarthritis would not completely affect these activities. Participants were questioned to score their capability of performing these tasks trichotomously, with either “able,” “need help,” or “unable” as choices for answers. We defined “disabled” participants as those who answered “need help” or “unable” to perform one or more items; they were defined as “non-disabled” otherwise, on the based on the previous studies.^26,27^

#### Motor functions

Their usual gait speed (UGS) was measured using a 5 m timed gait test. Participants were instructed to walk straight on an 11 m flat surface at their usual pace. A gait speed of 5 m was measured in the middle of the walkway.^28^ A goniometer measured the range of motion for knee extension and flexion,^29^ while a handheld dynamometer ^a^ assessed the isometric knee extension strength (IKES) to quantify knee muscle power.^30^ Range of motions and IKES were measured on both the operated and non-operated knees.

#### Pain status

The pain was evaluated by using the pain subscale of the Japanese Knee Injury and Osteoarthritis Outcome Score (KOOS-pain).^31^ Participants were asked about their pain in various situations one week before the evaluation date. Each question was scored in the range of 0–4. The normalized total score is converted to 100 points, with higher scores indicating less pain. KOOS-pain has satisfactory internal consistency (Cronbach’s alpha = 0.90) and correlates with the body pain subscale of the 36-item short-form health survey (*r* = 0.67, *p* < 0.01).^31^

#### Psychological factors

Depressive symptoms, pain catastrophizing, and self-efficacy were evaluated as potential psychological factors based on a previous systematic review.13,16 The Japanese version of the fifteen-item Geriatric Depression Scale (GDS-15) ^32^ was used to assess depressive symptoms. The GDS-15 has satisfactory internal consistency (Cronbach’s alpha = 0.83) and high detectability of depression (area under the curve of the receiver operating characteristic curve [ROC curve] = 0.96).^32^ The participants were questioned with “yes” or “no” answer choices. A total score of 0 to 15 was assigned, with higher scores indicating more depressive symptoms.

The Japanese version of the six-item short form of the Pain Catastrophizing Scale (PCS-6)^33^ was used to assess pain catastrophizing. PCS-6 had satisfactory internal consistency (Cronbach’s alpha = 0.90)^29^ and correlated to pain status (*r* = 0.30, *p* < 0.001).^34^ Participants were required to evaluate their experience of six emotions and thoughts during pain. Each item is evaluated on a scale of 0 (“not at all”) to 4 (“all the time”). A total score of 0 to 24 was assigned, with higher scores representing higher levels of catastrophe.

The Japanese version of the four-item short form of the Pain Self-Efficacy Questionnaire (PSEQ-4)^35^ was used to assess self-efficacy. The PSEQ-4 had satisfactory internal consistency (Cronbach’s alpha = 0.90) and correlated to pain status (*r* = −0.35, *p* < 0.001).^34^ The PSEQ-4 comprises of four questions measuring the participant’s confidence when conducting specific activities despite the pain. Each item was evaluated on a scale of 0 (“not at all confident”) to 6 (“completely confident”). The total score of 0 to 24 was assigned with higher scores representing more self-efficacy.

### Statistical analysis

#### Primary analyses

The participants were divided into two groups according to their IADL status six months after KA: the disabled (answered “need help” or “unable” for one or more items) and non-disabled (answered “able” for all items) groups. All preoperative baseline characteristics and measurements were compared between the two groups using the Mann–Whitney *U* test, chi-square, or student’s *t*-test. In the disabled group, the disability of each IADL tested at follow-up was recorded. Baseline characteristics were also compared between those who were followed-up and those who dropped out to assess the possibility of a selection bias.

Measurements with p < 0.05 in the two comparative groups were represented nominally based on the cutoff value for determining the presence of IADL disability at six postoperative months, calculated from the ROC curve. The ideal cutoff value for balancing the sensitivity and specificity of a test was determined as the point on the curve closest to (0, 1).^36^ Thereafter, logistic regression was used to predict the presence of IADL disability after six months of KA (dependent variable). The independent variables were included in each model separately, using nominal scales. In model 1, age, sex, KL-score of both knees, and operation type (TKA or UKA) were used as covariates. In model 2, the presence of IADL disability before the operation was added to model 1. KL-score on the non-operated side for participants with a history of TKA or UKA was treated as score 0.

#### Subgroup analyses

Additionally, participants were categorized into four subgroups based on pre-and postoperative IADL status: group 1 without IADL disability; group 2 with IADL disability before operation only; group 3 with IADL disability after operation only; and group 4 with IADL disability before and after the operation. UGS and KOOS pain were compared pre- and postoperatively in each group to confirm the treatment effect of KA using the paired *t*-test. The baseline characteristics in the four subgroups were also compared using the chi-square goodness of fit test, Fisher’s exact test, one-way analysis of variance followed by Tukey’s test, or Kruskal–Wallis test followed by the Steel–Dwass test. All data were analyzed using R version 4.0.3.^b^ A p-value < 0.05 was used to indicate statistical significance.

#### Sample size

In order to decide the sample size for logistic regression analysis, the number of cases (N) 10 k/p are needed; *k* is the number of independent variables as covariates, and *p* is defined as a ratio of responders to non-responders at the follow-up points.^37^ We predicted that eight factors out of all the variables would be required to avoid overfitting.^38^ In a previous report (n = 407), 60.9% of older adults with joint pain had an IADL disability.^26^ Based on this study, we hypothesized that the ratio *p* of non-disabled to disabled was 1:1.5 and calculated the minimum number of participants required for our study as 10 x 8/0.6 = 134.

## Results

### Study participants’ characteristics

Of the 220 participants who joined the study, 166 (75.5%) completed the follow-up assessment after surgery. Fifty-four (24.5%) participants dropped out, namely due to incomplete follow-up (n = 21), reparative surgery on the other knee within six months (n = 14), cancelation of the operation (n = 9), the onset of other diseases not related to surgery (n = 4), periprosthetic joint infection (n = 3), and periprosthetic fracture (n = 3) (Table 1). Baseline characteristics were not significantly different between those who were followed up and dropped out.

**Table 1.**
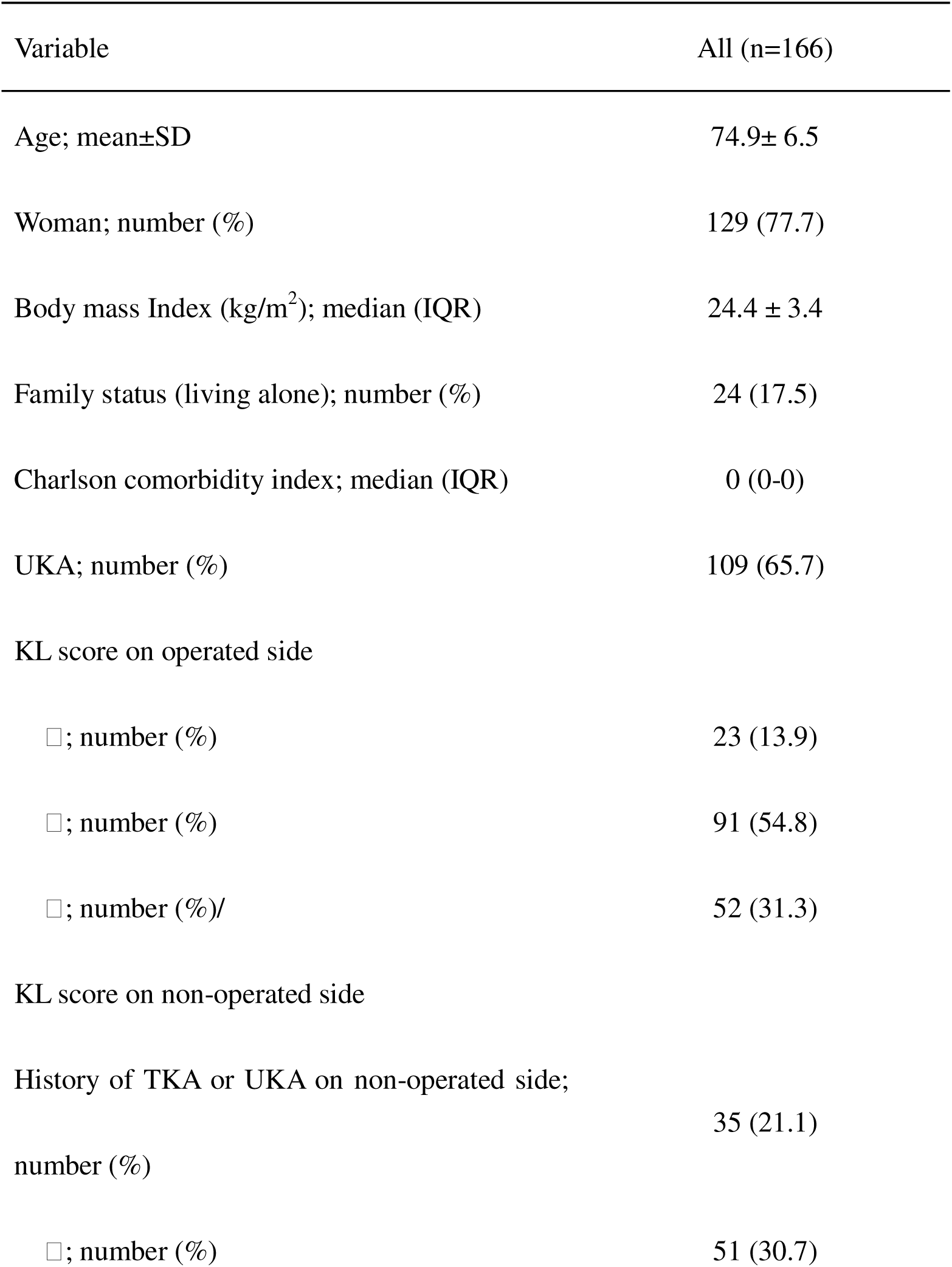
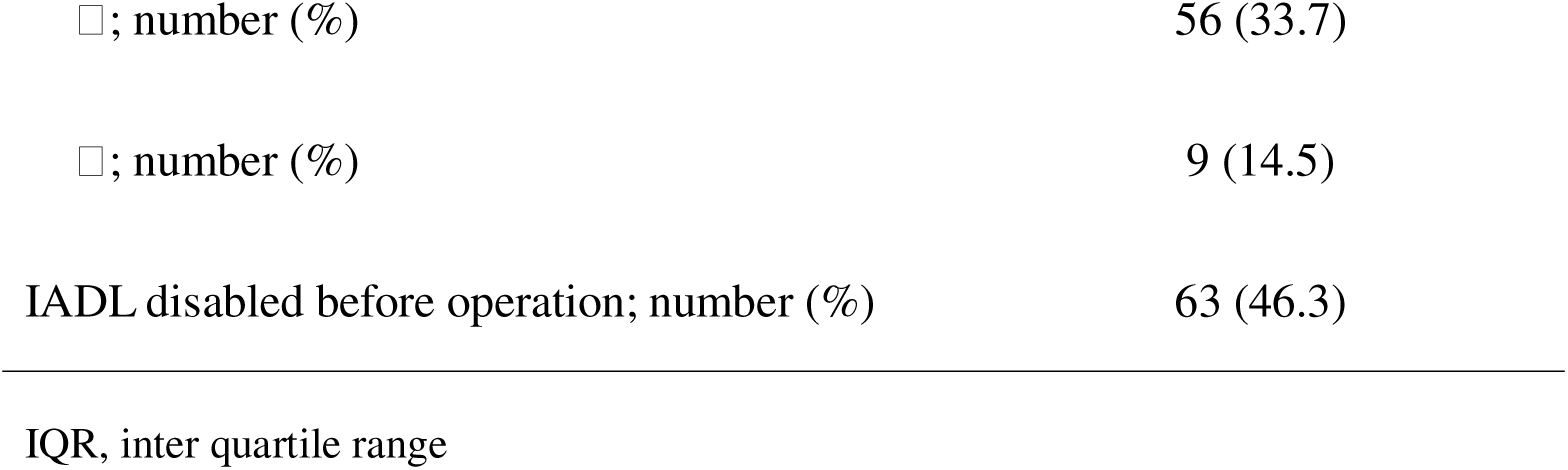
Baseline characteristics of the study population

### Preoperative predictors of postoperative IADL disability

Of the participants who were followed up, 83 (50.0%) had IADL disability six months after surgery. Sixty-one (69.3%) struggled with shopping, and 47 (53.4%) struggled using transportation (Table 2). The non-disabled group was significantly younger (p = 0.004), with a larger percentage of women (85.5% vs. 69.9%; p = 0.03) than the IADL-disabled group. The percentage of patients showing IADL disability before surgery was 20.5% and 72.3% (p < 0.001) in the non-disabled and disabled groups, respectively. In the IADL non-disabled group, the values of UGS, IKES (on the non-operated side), and PSEQ-4 were significantly higher (Table 3). Based on the two groups’ comparison, the cutoff values of UGS, IKES (on the non-operated side), and PSEQ-4 were calculated by inspecting the ROC curve to discern the presence of IADL disability at six months after surgery (Figure 1). The optimal cutoff values are shown in Table 4. UGS was a significant independent variable in models 1 and 2, and IKES (on the non-operated side) and PSEQ-4 were significant in model 1 but not in model 2. (Table 5)

**Figure 1.**
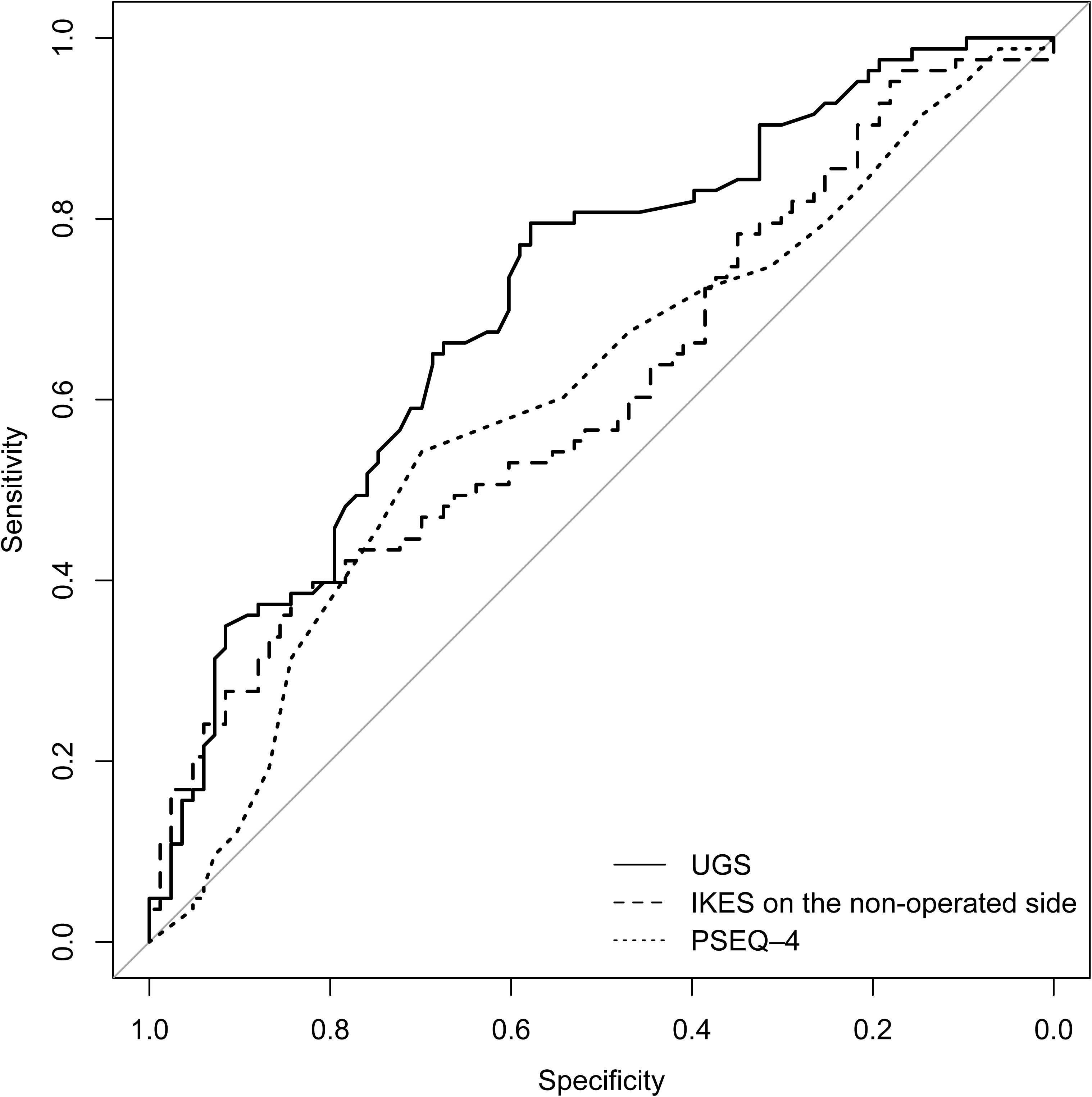
The ROC-curves of UGS, IKES (on the non-operated side), and PSEQ-4

**Table 2.** Content of disability in the IADL-disabled group

| Items | IADL disabled 6-months after operation (n=83)* |
| --- | --- |
| Preparing food | 33 (37.5) |
| Shopping | 61 (69.3) |
| Housekeeping | 40 (45.5) |
| Doing laundry | 6 (6.8) |
| Using transportation | 47 (53.4) |
| Handling finances | 15 (17.0) |

\* Number (%)

**Table 3.**
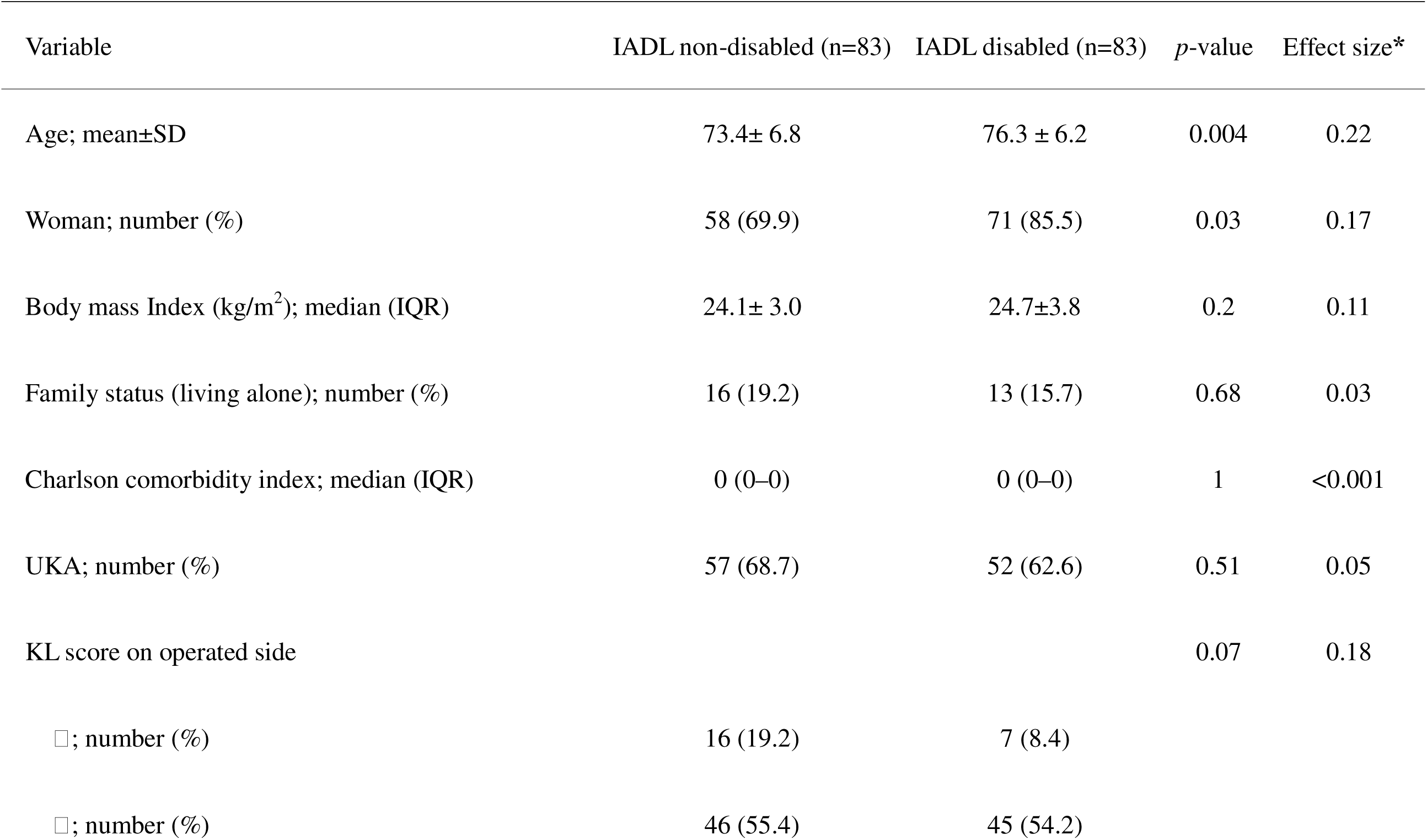
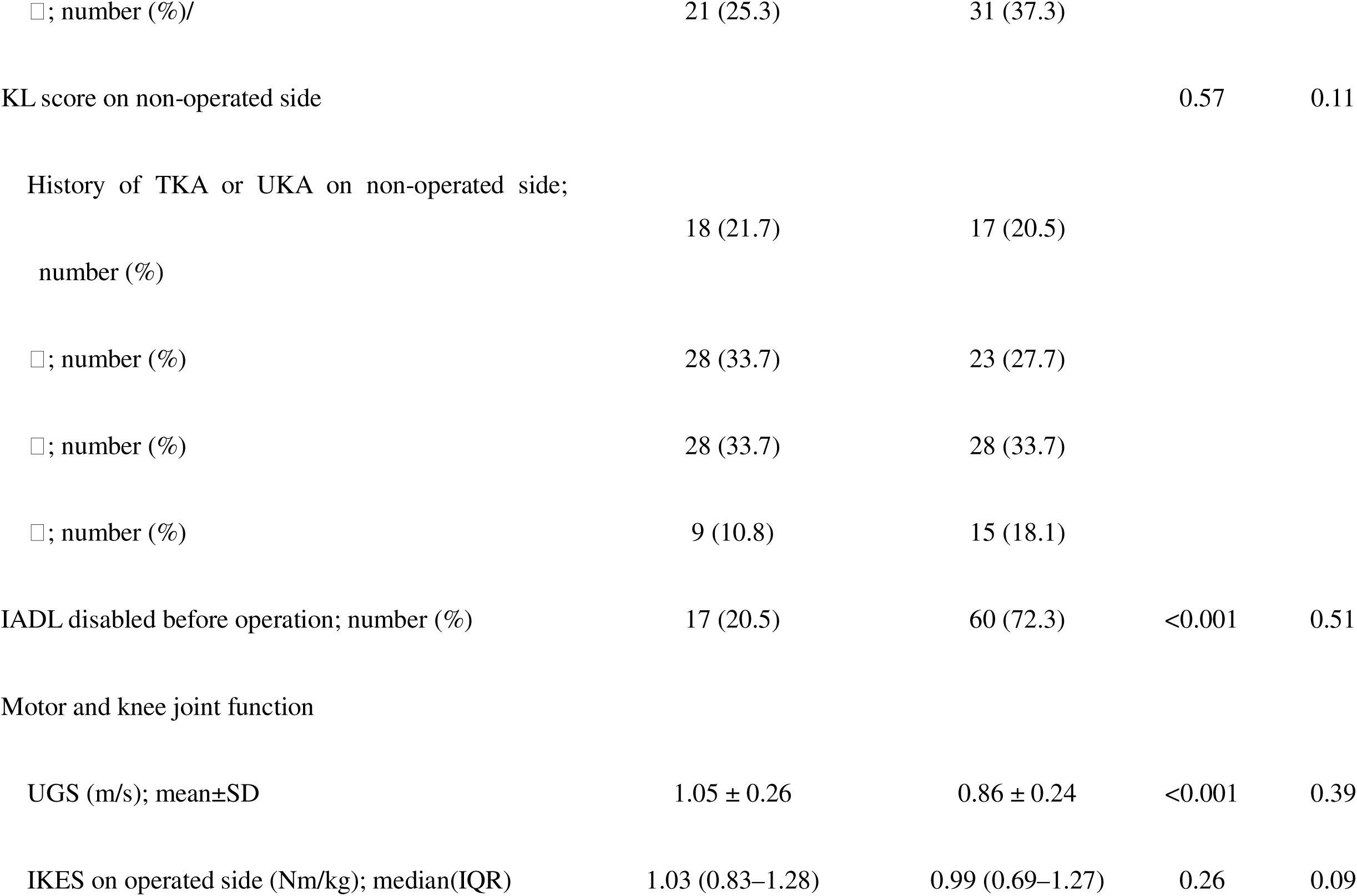
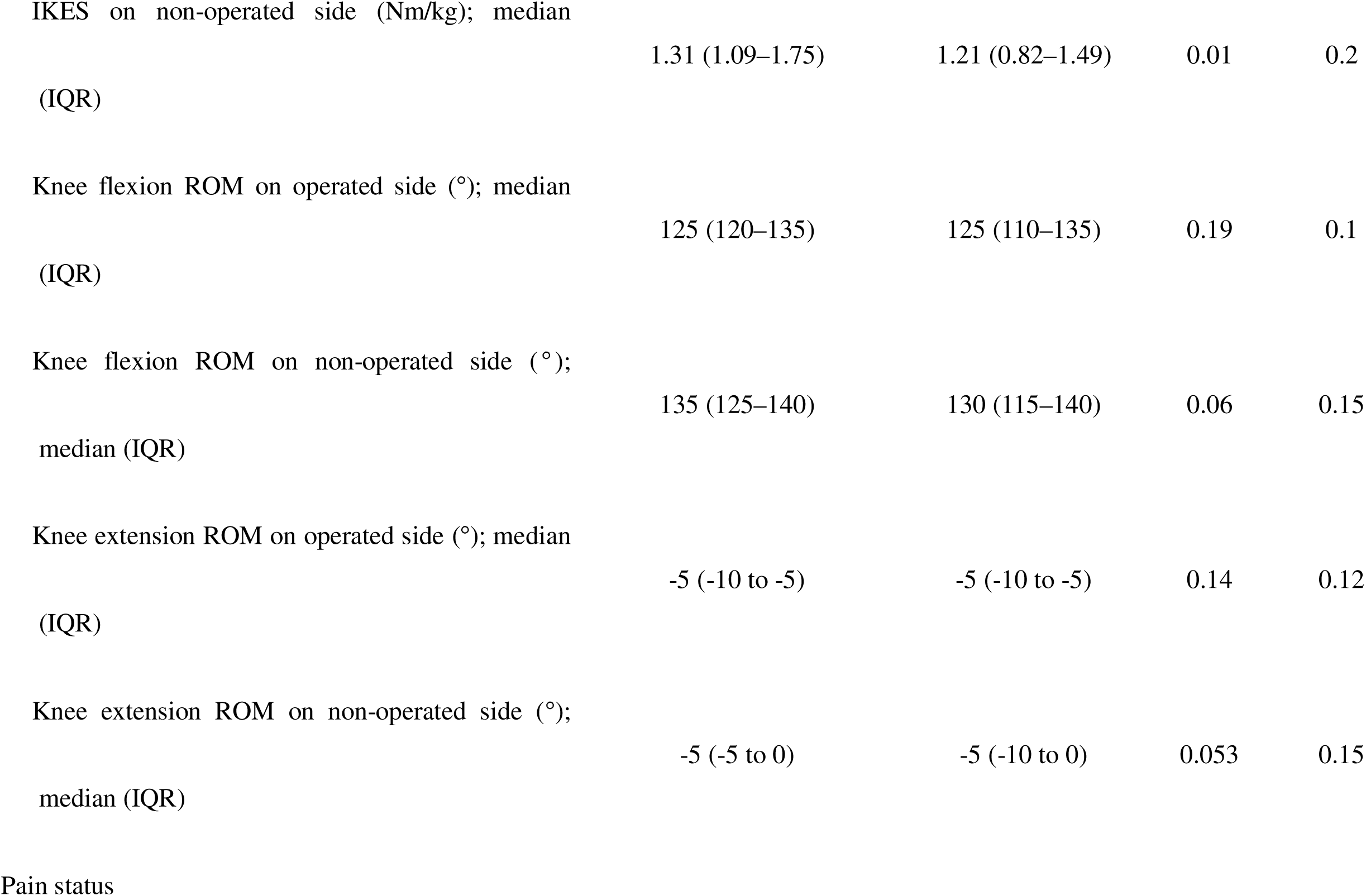
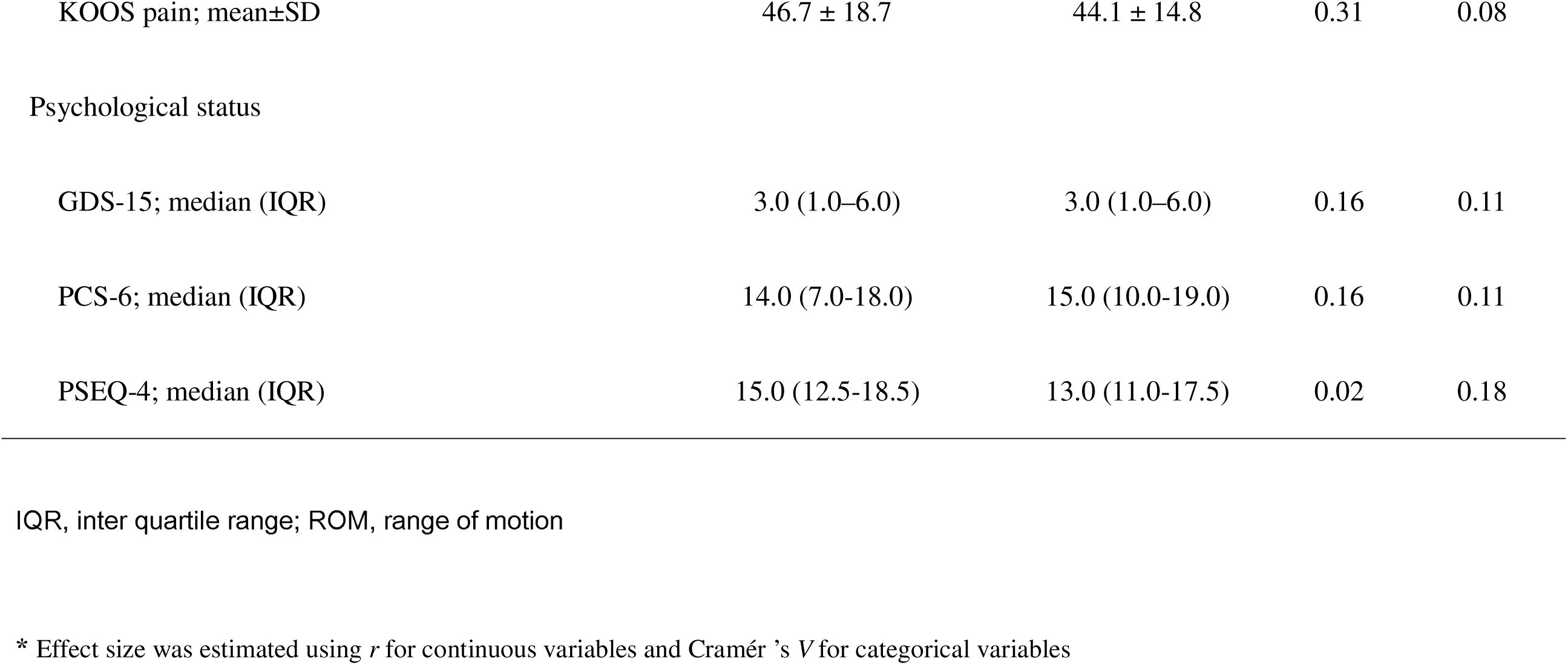
Comparison of baseline characteristics and preoperative measurements between the IADL non-disabled and disabled groups

**Table 4.** Variable characteristics to discriminate those with or without IADL disability six months after KA

| Variables | Cut off | AUC (95%CI) | Sensitivity (%) | Specificity (%) | OR (95% CI) |
| --- | --- | --- | --- | --- | --- |
| UGS (m/s) | 1.0 | 0.71 (0.63–0.79) | 57.8 | 79.5 | 5.32 (2.57–10.6) |
| IKES on the non-operated side (Nm/kg) | 1.0 | 0.61 (0.53–0.70) | 84.3 | 38.6 | 3.38 (1.61–7.07) |
| PSEQ–4 (points) | 13 | 0.60 (0.52–0.69)) | 69.9 | 54.2 | 2.49 (1.29–4.81) |

AUC, area under the curve; CI, confidence interval; OR, odds ratio

**Table 5.**
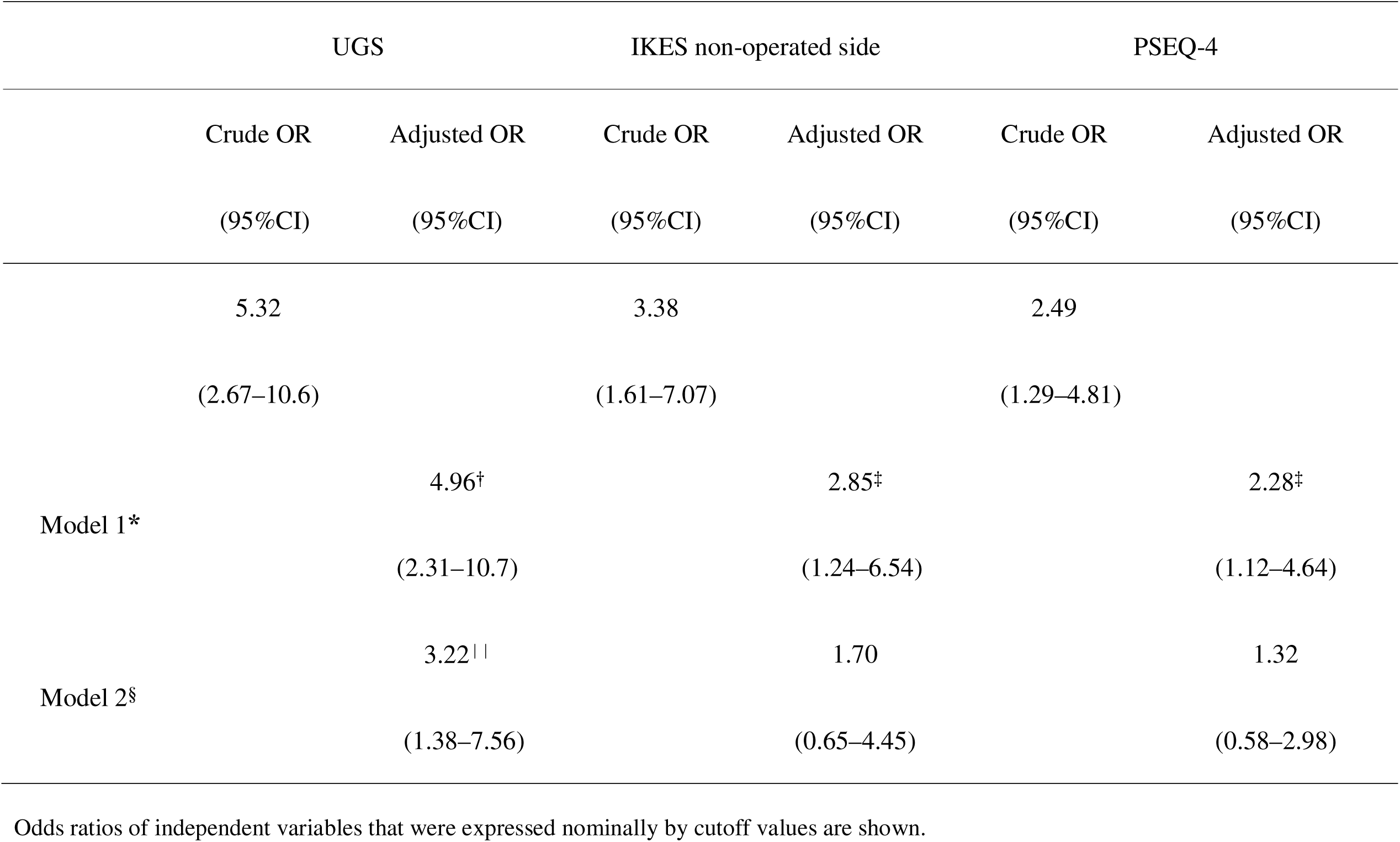
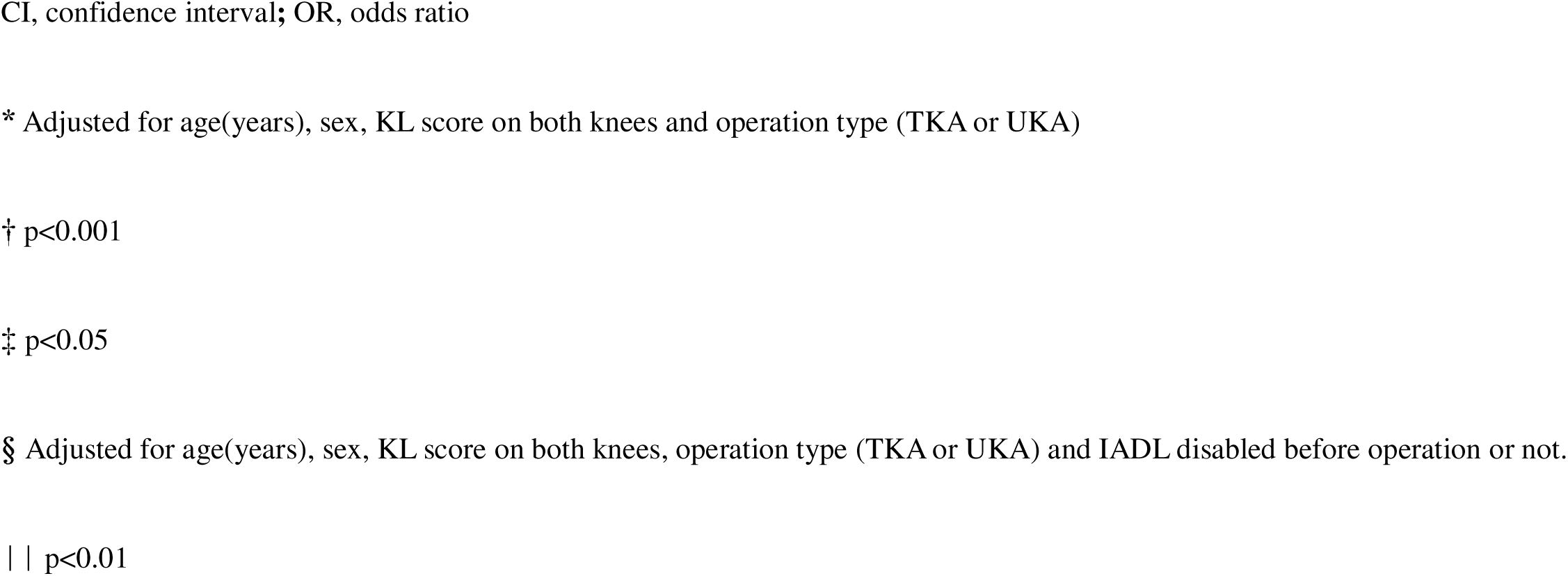
ORs of independent variables in a logistic regression model with IADL disability status at six months after KA as dependent variable

|  | UGS |  | IKES non-operated side |  | PSEQ-4 |  |
| --- | --- | --- | --- | --- | --- | --- |
|  | Crude OR | Adjusted OR | Crude OR | Adjusted OR | Crude OR | Adjusted OR |
|  | (95%CI) | (95%CI) | (95%CI) | (95%CI) | (95%CI) | (95%CI) |
|  | 5.32 |  | 3.38 |  | 2.49 |  |
|  | (2.67–10.6) |  | (1.61–7.07) |  | (1.29–4.81) |  |
| Model 1* |  | 4.96<sup>†</sup> |  | 2.85<sup>‡</sup> |  | 2.28<sup>‡</sup> |
|  |  | (2.31–10.7) |  | (1.24–6.54) |  | (1.12–4.64) |
| Model 2<sup>§</sup> |  | 3.22 |  | 1.70 |  | 1.32 |
|  |  | (1.38–7.56) |  | (0.65–4.45) |  | (0.58–2.98) |

Odds ratios of independent variables that were expressed nominally by cutoff values are shown.
CI, confidence interval; OR, odds ratio
\* Adjusted for age(years), sex, KL score on both knees and operation type (TKA or UKA)
† $p < 0.001$
‡ $p < 0.05$
§ Adjusted for age(years), sex, KL score on both knees, operation type (TKA or UKA) and IADL disabled before operation or not.
|| $p < 0.01$

### Comparison of the treatment effect of KA among subgroups

In the baseline characteristics, comparisons among subgroups, significant differences in the age, and number of women were detected. Regarding KL-score (on the non-operated side), there was a cell with 0 frequency, making statistical analysis impossible (Table 6). KOOS-pain values were significantly higher in all subgroups after the operation than before. The values of UGS after the operation were significantly higher than before, the group 3 being the exception (Table 7).

**Table 6.**
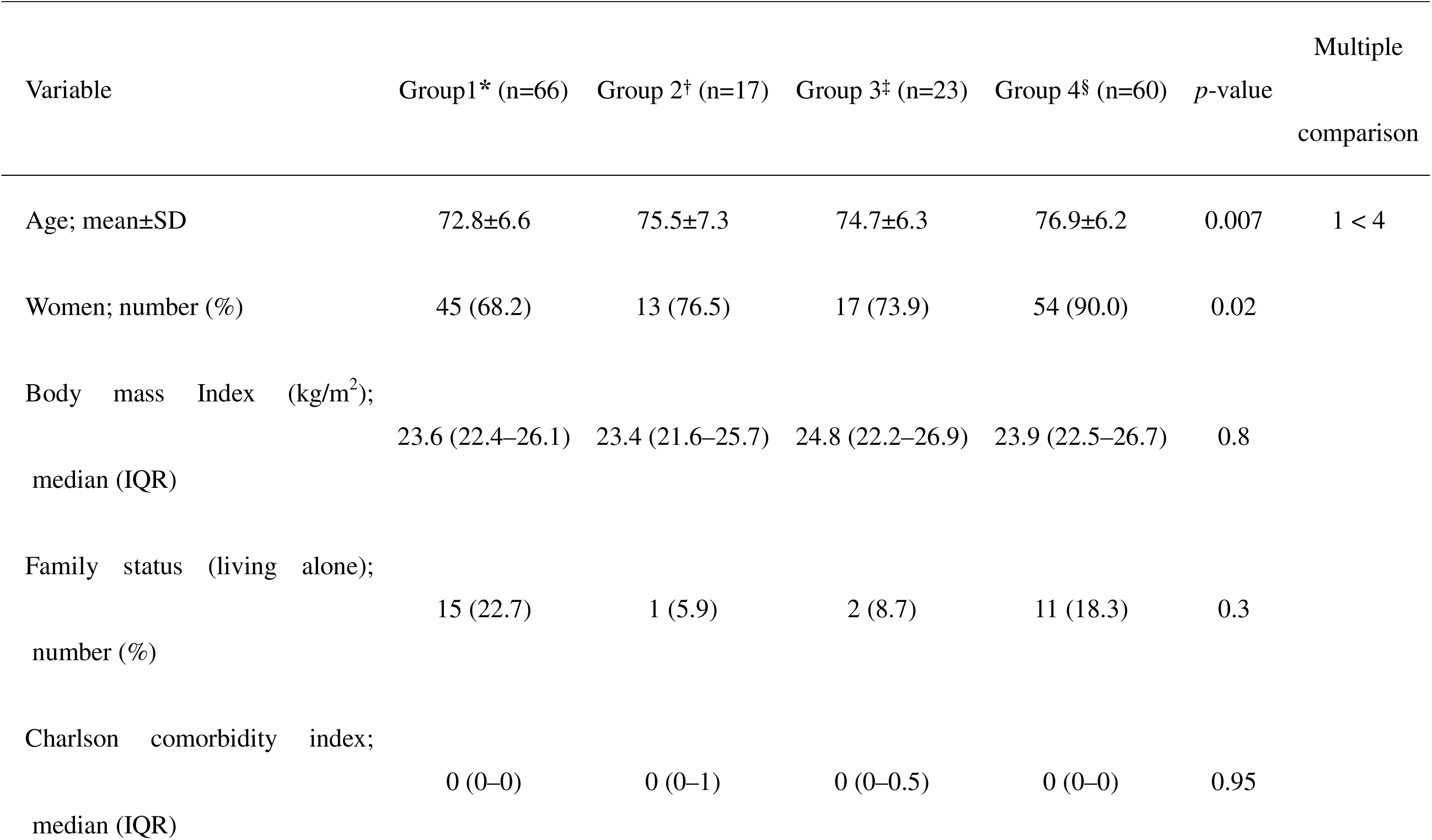
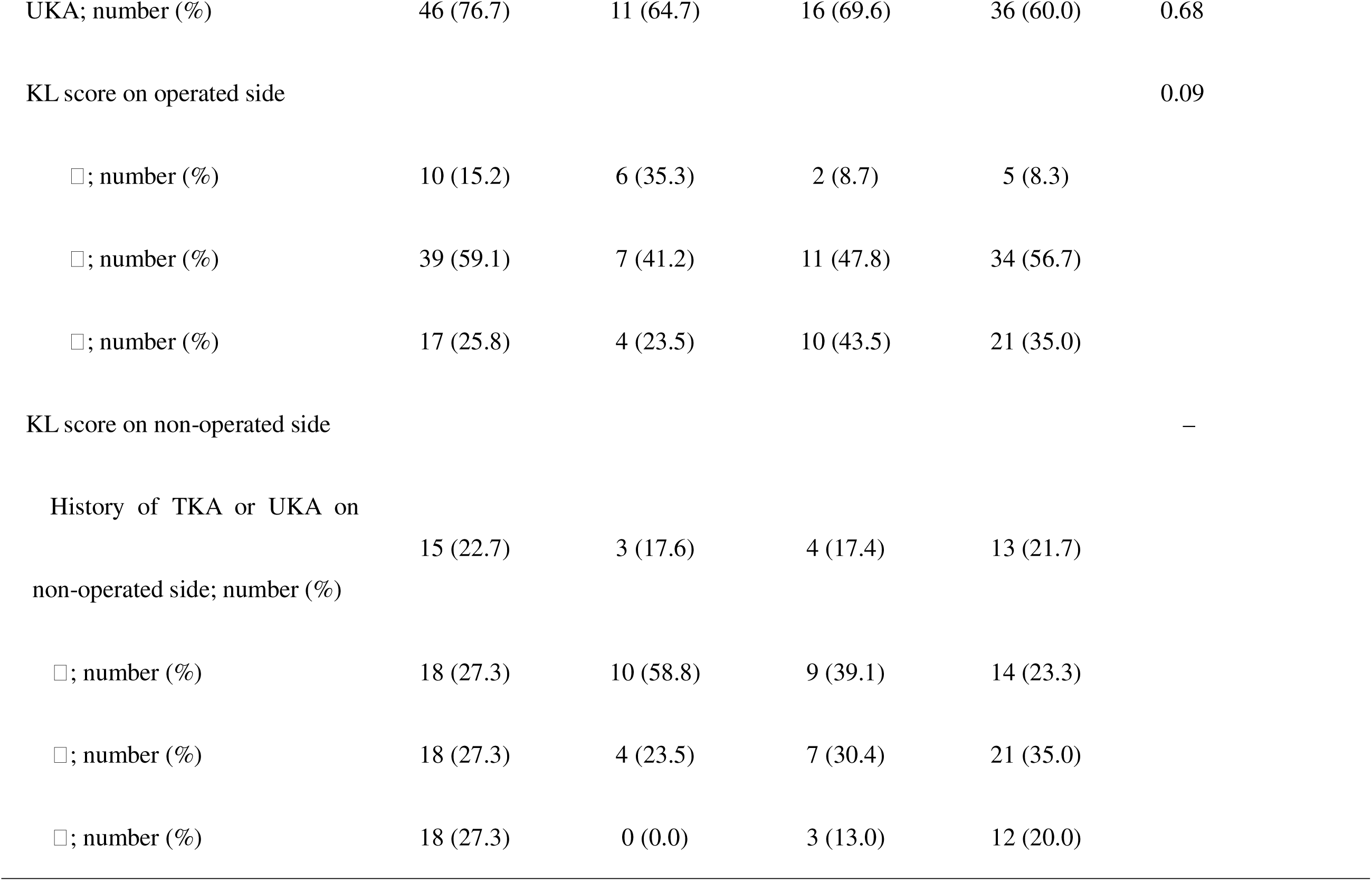
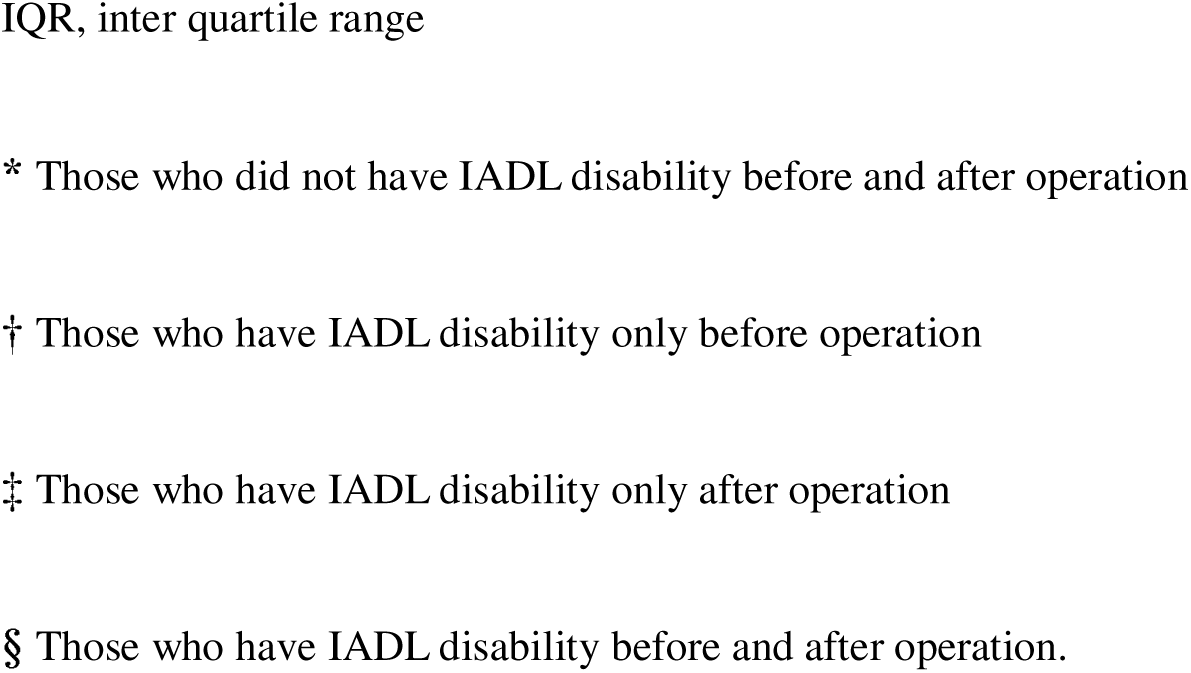
Demographic comparison in four subgroups

| Variable | Group1* (n=66) | Group 2† (n=17) | Group 3‡ (n=23) | Group 4§ (n=60) | *p*-value | Multiple<br>comparison |
| --- | --- | --- | --- | --- | --- | --- |
| Age; mean±SD | 72.8±6.6 | 75.5±7.3 | 74.7±6.3 | 76.9±6.2 | 0.007 | 1 < 4 |
| Women; number (%) | 45 (68.2) | 13 (76.5) | 17 (73.9) | 54 (90.0) | 0.02 |  |
| Body mass Index (kg/m<sup>2</sup>);<br>median (IQR) | 23.6 (22.4–26.1) | 23.4 (21.6–25.7) | 24.8 (22.2–26.9) | 23.9 (22.5–26.7) | 0.8 |  |
| Family status (living alone);<br>number (%) | 15 (22.7) | 1 (5.9) | 2 (8.7) | 11 (18.3) | 0.3 |  |
| Charlson comorbidity index;<br>median (IQR) | 0 (0–0) | 0 (0–1) | 0 (0–0.5) | 0 (0–0) | 0.95 |  |

|  |  |  |  |  |  |
| --- | --- | --- | --- | --- | --- |
| UKA; number (%) | 46 (76.7) | 11 (64.7) | 16 (69.6) | 36 (60.0) | 0.68 |
| KL score on operated side |  |  |  |  | 0.09 |
| □; number (%) | 10 (15.2) | 6 (35.3) | 2 (8.7) | 5 (8.3) |  |
| □; number (%) | 39 (59.1) | 7 (41.2) | 11 (47.8) | 34 (56.7) |  |
| □; number (%) | 17 (25.8) | 4 (23.5) | 10 (43.5) | 21 (35.0) |  |
| KL score on non-operated side |  |  |  |  | — |
| History of TKA or UKA on<br>non-operated side; number (%) | 15 (22.7) | 3 (17.6) | 4 (17.4) | 13 (21.7) |  |
| □; number (%) | 18 (27.3) | 10 (58.8) | 9 (39.1) | 14 (23.3) |  |
| □; number (%) | 18 (27.3) | 4 (23.5) | 7 (30.4) | 21 (35.0) |  |
| □; number (%) | 18 (27.3) | 0 (0.0) | 3 (13.0) | 12 (20.0) |  |

IQR, inter quartile range
\* Those who did not have IADL disability before and after operation
† Those who have IADL disability only before operation
‡ Those who have IADL disability only after operation
§ Those who have IADL disability before and after operation.

**Table 7.**
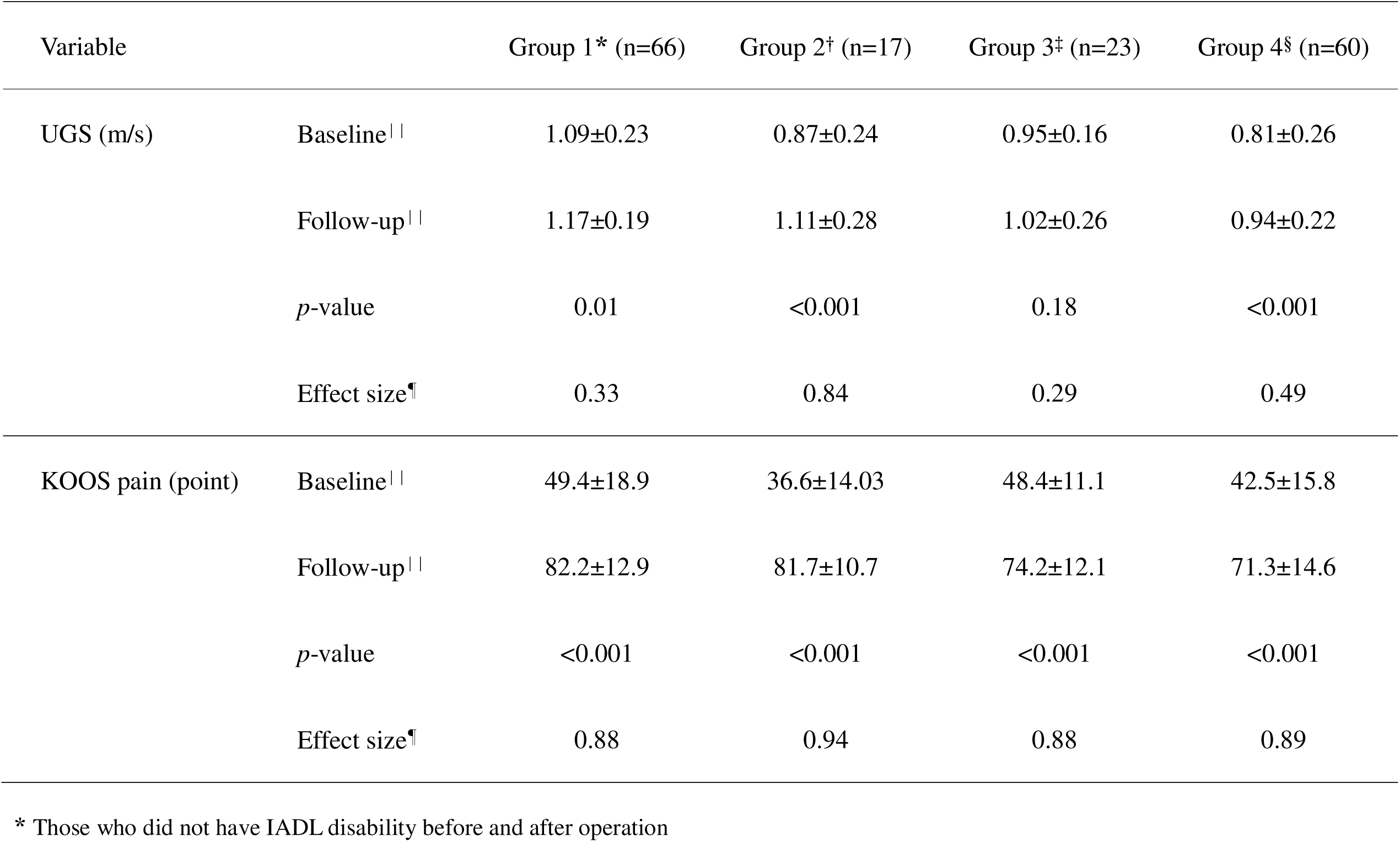
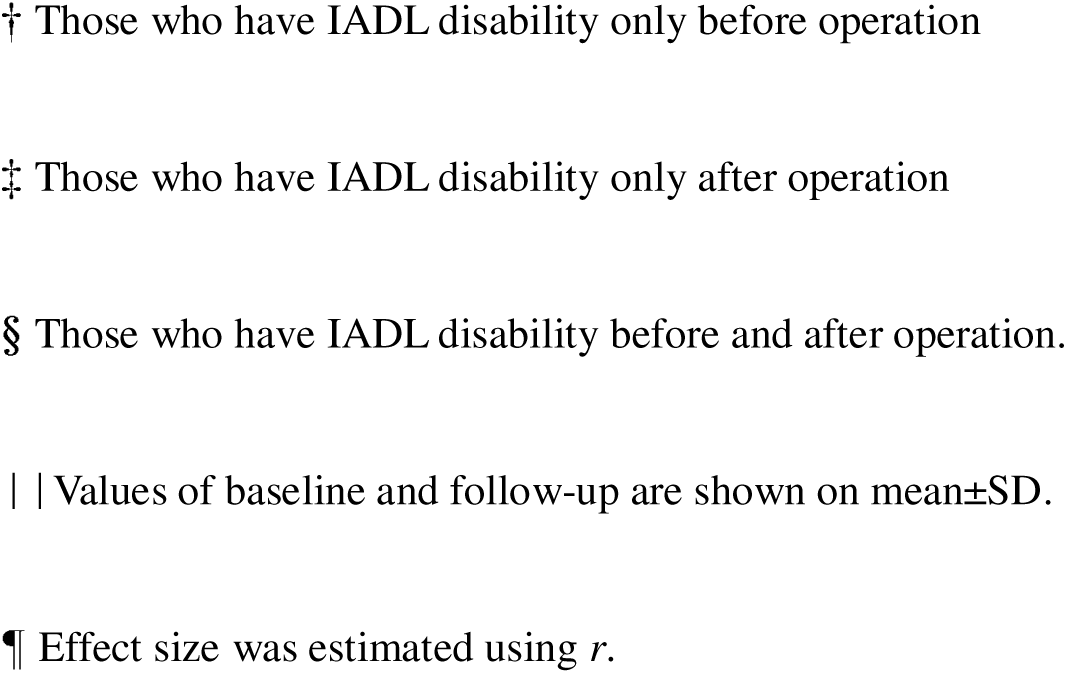
Comparison of KOOS-pain and UGS between pre- and postoperative subgroups

| Variable |  | Group 1* (n=66) | Group 2<sup>†</sup> (n=17) | Group 3<sup>‡</sup> (n=23) | Group 4<sup>§</sup> (n=60) |
| --- | --- | --- | --- | --- | --- |
| UGS (m/s) | Baseline | 1.09±0.23 | 0.87±0.24 | 0.95±0.16 | 0.81±0.26 |
|  | Follow-up | 1.17±0.19 | 1.11±0.28 | 1.02±0.26 | 0.94±0.22 |
|  | *p*-value | 0.01 | <0.001 | 0.18 | <0.001 |
|  | Effect size<sup>¶</sup> | 0.33 | 0.84 | 0.29 | 0.49 |
| KOOS pain (point) | Baseline | 49.4±18.9 | 36.6±14.03 | 48.4±11.1 | 42.5±15.8 |
|  | Follow-up | 82.2±12.9 | 81.7±10.7 | 74.2±12.1 | 71.3±14.6 |
|  | *p*-value | <0.001 | <0.001 | <0.001 | <0.001 |
|  | Effect size<sup>¶</sup> | 0.88 | 0.94 | 0.88 | 0.89 |

\* Those who did not have IADL disability before and after operation
† Those who have IADL disability only before operation
‡ Those who have IADL disability only after operation
§ Those who have IADL disability before and after operation.
| | Values of baseline and follow-up are shown on mean ± SD.
¶ Effect size was estimated using *r*.

## Discussion

This study investigated preoperative predictors of IADL disability in older adults after six months of KA, where half of the participants reported disabilities. The logistic regression analysis adjusted for covariates showed that only gait speed was a valid predictor of IADL disability six months after KA. Thus, the results of this study partially supported our hypothesis.

### Preoperative predictors of postoperative IADL disability

Our study suggested that the cutoff value for preoperative UGS was 1.0 m/s. The area under the curve needed to be at least 0.7 to be considered satisfactory.^36^ Thus, our cutoff value for preoperative UGS exceeded the minimum criterion. A previous prospective study in older adults showed that participants with UGS below 1.0 m/s had a sharper increase in the 2-year incidence of IADL disability.^39^ Other cohort studies involving older adults have also revealed a 1.0 m/s cutoff value for risk prediction of health events.^40,41^ Considering the incidence of BADL disability, 0.8 m/s has been suggested as a more sensible cutoff value.^42^ Therefore, considering the hierarchical relationship between IADL and BADL,^9^ our cutoff value for UGS was valid.

The predictive power of PSEQ-4 and IKES (on the non-operated side) was low in this study. However, previous studies showed that self-efficacy^13^ and muscle strength^14^ were predictors of decreased functional status after KA. Older adults with lesser self-efficacy consistently limit their involvement in activities,^43^ suggesting that self-efficacy influences activities independently of motor functions. Thus, these factors should not be overlooked when considering disability after surgery.

George et al. have shown that the number of older adults with preoperative IADL disability improved after TKA.^20^ However, the proportion of older adults in our study who presented with IADL disability before the operation worsened after TKA or UKA. The aforementioned study assessed fewer IADLs and included younger participants than those in our study, which we believe could have contributed to the different results.

Our participants’ primary IADL disability items were outdoor activities, such as shopping (69.3%) and using transportation (53.4%), which require gait proficiency. When comparing subgroups, only those presenting with new IADL disability after the operation (group 3) failed to improve UGS after KA. In group 4, UGS significantly improved after KA despite having the lowest baseline and follow-up values of the four groups. As a result, we believe that those with a slow preoperative gait speed, which does not increase to a certain value postoperatively, tend to present IADL disability six months after KA, even if their pain status has improved.

Some studies have shown that preoperative functional limitation based on a total score calculated from subjective difficulty in various activities affects postoperative disability.^44–46^ Thus, capacities of individual activities such as gait or stair climbing are unknown. The strength of our findings lies in the predictive power of preoperative gait speed as a modifiable factor. Additionally, the specific value of UGS, which can be measured easily and objectively, was suggested as a promising preoperative predictor. Based on the clinical practice guideline, older patients awaiting KA are recommended preoperative exercise programs to improve strengthening, flexibility, and functions before surgery.^17^ However, the specific regimen or goal for exercise needs to be clarified. Our results could be helpful for preoperative planning and intervention. In particular, improving preoperative gait speed may enhance the IADL status, and our cut-off value can be one of the goals for preoperative exercise.

### Comparison of treatment effect of KA among subgroups

The systematic review by Sayah et al. revealed that functional limitations, including IADL disability, improved drastically in six months and gradually kept improving 2 years after TKA.^47^ Therefore, our study population, especially participants in group 3, could potentially improve IADL disability six months postoperatively. However, general rehabilitative intervention periods after KA have been set within six months.^18,19^ Additionally, previous cohort studies have shown that the presence of even one IADL disability affected all-cause mortality^10^ and the cognitive function decline rate.^11^ Therefore, as implications of our study, patients walking slowly before surgery or unable to improve gait speed sufficiently after surgery are at high risk for adverse events and require frequent follow-up even beyond six months postoperatively. Furthermore, a physical therapist should be consulted beyond general rehabilitative intervention periods.

### Study limitations

The present study had some limitations. First, we could not determine how preoperative factors affected IADL disability after KA. Second, ours was a single-center analysis; the area distribution where the participants lived was limited. Third, our study population included those with a history of KA, and we could not exclude the effect of past KA on IADL disability. However, the number of those with a history of TKA or UKA was similar between IADL disabled and IADL non-disabled groups.

## Conclusion

Our study revealed the importance of evaluating UGS before a KA as a predictor of six-month postoperative IADL disability in older adults, in addition to preoperative IADL status. Those with poor mobility before surgery should be carefully rehabilitated after KA. Future studies investigating IADL disability during long-term follow-up periods must further validate our findings.

### Suppliers list

a: μ-tus F-1, Anima Corporation, 3-65-1, Shimoishihara, Chofu, Tokyo, 182-0034, Japan

b: R version 4.0.3, R Foundation for Statistical Computing, Welthandelsplatz 1, 1020 Vienna, Austria

## Data Availability

All data produced in the present study are available upon reasonable request to the authors

## Acknowledgements

The authors thank PTs. D. Kurihara, K. Imahira, K. Suda and G. Tamura for their cooperation with data collection.

## Device Status Statement

The manuscript submitted does not contain information about medical device.

## Data availability statement

The data that support the findings of this study are available from the corresponding author, Nanjo, K., upon reasonable request.

## Abbreviations

BADL: basic activities of daily living
GDS-15: fifteen-item Geriatric Depression Scale
IADL: instrumental activities of daily living
IKES: isometric knee extension strength
KA: knee arthroplasty
KL-score: Kellgren-Lawllence score
KOOS-pain: pain subscale of the knee injury and osteoarthritis outcome score
PCS-6: six-item short form of the pain catastrophizing scale
PSEQ-4: four-item short form of the pain self-efficacy questionnaire
ROC curve: receiver characteristic operating curve
TKA: total knee arthroplasty
UGS: usual gait speed
UKA: unicompartmental knee arthroplasty

